# Global and local mobility as a barometer for COVID-19 dynamics

**DOI:** 10.1101/2020.06.13.20130658

**Authors:** Kevin Linka, Alain Goriely, Ellen Kuhl

## Abstract

The spreading of infectious diseases including COVID-19 depends on human interactions. In an environment where behavioral patterns and physical contacts are constantly evolving according to new governmental regulations, measuring these interactions is a major challenge. Mobility has emerged as an indicator for human activity and, implicitly, for human interactions. Here we study the coupling between mobility and COVID-19 dynamics and show that variations in global air traffic and local driving mobility can be used to stratify different disease phases. For ten European countries, our study shows maximal correlation between driving mobility and disease dynamics with a time lag of 14.6 ± 5.6 days. Our findings suggests that local mobility can serve as a quantitative metric to forecast future reproduction numbers and identify the stages of the pandemic when mobility and reproduction become decorrelated.

## Introduction

A barometer is an instrument that measures air pressure. Its main purpose is to forecast short term changes in the weather. It is often imprecise as it relies on a number of assumptions linking variations of air pressure to atmospheric conditions. Nonetheless, overall, we accept it as an important tool in weather prediction that has saved countless lives since its introduction for forecasting in the 19th century. In an evolving crisis like the COVID-19 pandemic, the world is in a dire need of a barometer that would provide us with reliable estimates of the disease dynamics when lockdown measures are either enacted or removed.

The transmission of an infectious disease depends mostly on three key aspects: the biology of the disease when individuals are in contact, social and human behavior that dictate the way individuals interact, and the physics of such contacts (closed spaces, close contact, crowded spaces) (*1*). We still do not know about the details of the COVID-19 pathology, and in particular how different age or population groups either transmit or are susceptible to the disease (*2,3*). But, the disease pathogenicity and transmissibility cannot be altered in the absence of a vaccine or preventive treatments. On the behavioral side, there has been a dramatic change in everyday habits with widespread adoption of new rules to prevent both close contact between individuals and the exchange of contaminated bodily fluids. The remaining factors, frequency and type of contact, depend on human activity. Work, school, and leisure inevitably increase the number of contacts, and hence the risk of transmission that occurs in everyday life (*4*). At the global level, human mobility, tracked by mobile phone use, has recently emerged as a possible proxy for human activity. Studies of the effective reproduction number *R*(*t*), the average number of secondary infections caused by an infected person, against in-state human mobility data in China (*5-7*) and in the United States (*8,9*) have demonstrated that as long as the epidemic is ongoing, mobility and reproduction number are indeed correlated (*10*). Interestingly, successful exit strategies reveal a decorrelation between these mobility and reproduction which implies that an increase in activity does not lead to an increase in infection.

When considered together, current mobility data and disease indicators follow the typical pattern shown in Fig. 1(a) with four distinct phases: Phase I of exponential growth during the initial outbreak of the disease; Phase II of outbreak control during which global and local mobility are rapidly reduced; Phase III of reduced growth under lockdown with reduced local and global mobility; and Phase IV of gradual exit during which lockdown measures are successively released and mobility increases while the number of new cases continues to decrease. The end of Phase IV marks the beginning of the second wave with newly increasing case numbers. An important feature of the outbreak dynamics is that a reduction in the number of new cases is delayed by about two weeks compared to the reduction in mobility and that this feature appears systematically in across countries. This is not particularly surprising as it is the primary goal of lockdown measures. However, we will show that another key property of the data is the high correlation between mobility data and the reproduction number *after* lockdown. We can formally exploit this property to understand the effects of human activity on the reproduction number.

**Figure 1:**
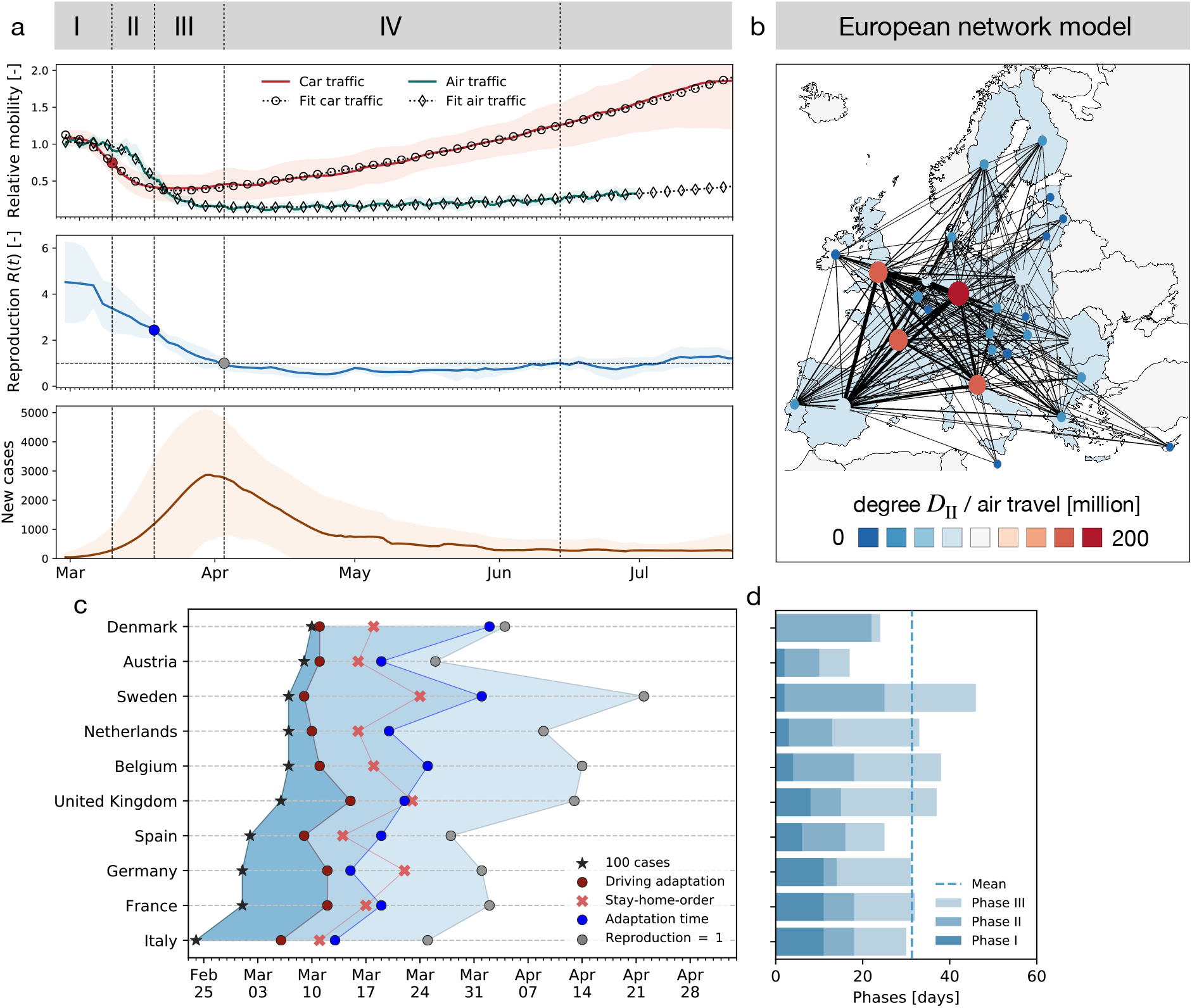
Phases of the COVID-19 outbreak in terms of of global and local mobility, reproduction number, and reported cases. (a) Phase I: Exponential growth during initial disease outbreak; Phase II: Outbreak control with rapidly reduced global and local mobility; Phase III: Reduced growth under lockdown with reduced local and global mobility; Phase IV: Gradual exit with successively released lockdown measures, increased mobility, decreasing number of new cases, and low effective reproduction number, indicating that many behavioral changes are still in place. Solid lines represent means values erf all European countries, shaded areas their standard deviations; (b) Global mobility network of European countries with 26 nodes and the 201 most traveled edges; (c) Phase evolution in 10 European countries; (d) Duration of individual phases: Dashed line indicates mean duration of 31 ± 8 days to reach *R*(*t*) = 1; Phase I with duration of 6 ± 4 days, Phase II with 11 ± 7 days and Phase III with 14 ± 7 days.

Modeling has proven to be a key element to understand disease dynamics (*11,12*) and establish new public health policies (*13*). While most models rely on standard population models such as the SEIR model, there are many refinements and variations that take into account contact between different groups, susceptibility, the effect of air travel through networks, and the effect of testing and lockdown (*3,14*). These models can be deterministic, representing average quantities, or stochastic, associated with uncertainties in data and disease dynamics. Due to the large uncertainties associated with model parameters, most of the actual work consists in devising appropridte statistical methods to infer parameters from incomplete data. This process naturally generates confidence intervals for any prediction based on these models. Precise predictions require more fine-grained models (*15*). These models consider multiple effects and mirror the complexity of interaction in societies; they include many parameters, including mobility and proximity data, that are difficult to track, especially since rapid changes in behavior often render a-few-day-old studies and historic trends irrelevant.

A complementary approach is to consider coarse-grained models with fewer parameters. While these models cannot be used to make precise long-term predictions of the number of infections or deaths on a given day, they are particularly valuable when predicting global trends in a robust manner since they only rely on a few parameters that are almost entirely determined by the data alone. Here we adopt this approach to study the relation between mobility and disease dynamics. We use the well-established Susceptible - Exposed - Infectious - Recovered SEIR compartment model with a network structure and evaluate the local disease dynamics at the level of a node, which represents an individual country. We incorporate the local driving mobility at the nodal level and the global mobility at the network level through passenger air travel with diffusive transport. To take full advantage of prior knowledge about the model parameters and the available data of each country, we combine our network model with a hierarchical Bayesian parameter inference and apply Markov Chain Monte Carlo sampling to obtain posterior distributions to our model uncertainties.

## Results

### Reduced mobility drastically reduces the effective reproduction number

Figure 2 shows the relative change in air and car traffic with respect to baseline before the outbreak in early February 2020. Each dot represents the day at which the corresponding country reached 100 or more cases. We notice that different countries share the same overall dynamics: an initial plateau followed by a sudden drop in air traffic and a rapid drop in car traffic followed by a gradual increase. The data reveal three interesting trends: First, local mobility decreases before global mobility with minima in the week of March 17-24 for car and March 24-31 for air traffic. Second, many countries experience a dramatic reduction of mobility, it seems almost voluntary, *before* the national outbreak becomes apparent. Third, there is a clear South-North divide in reduction of local mobility with Spain and Italy experiencing the largest reduction and Sweden and Finland the smallest.

**Figure 2:**
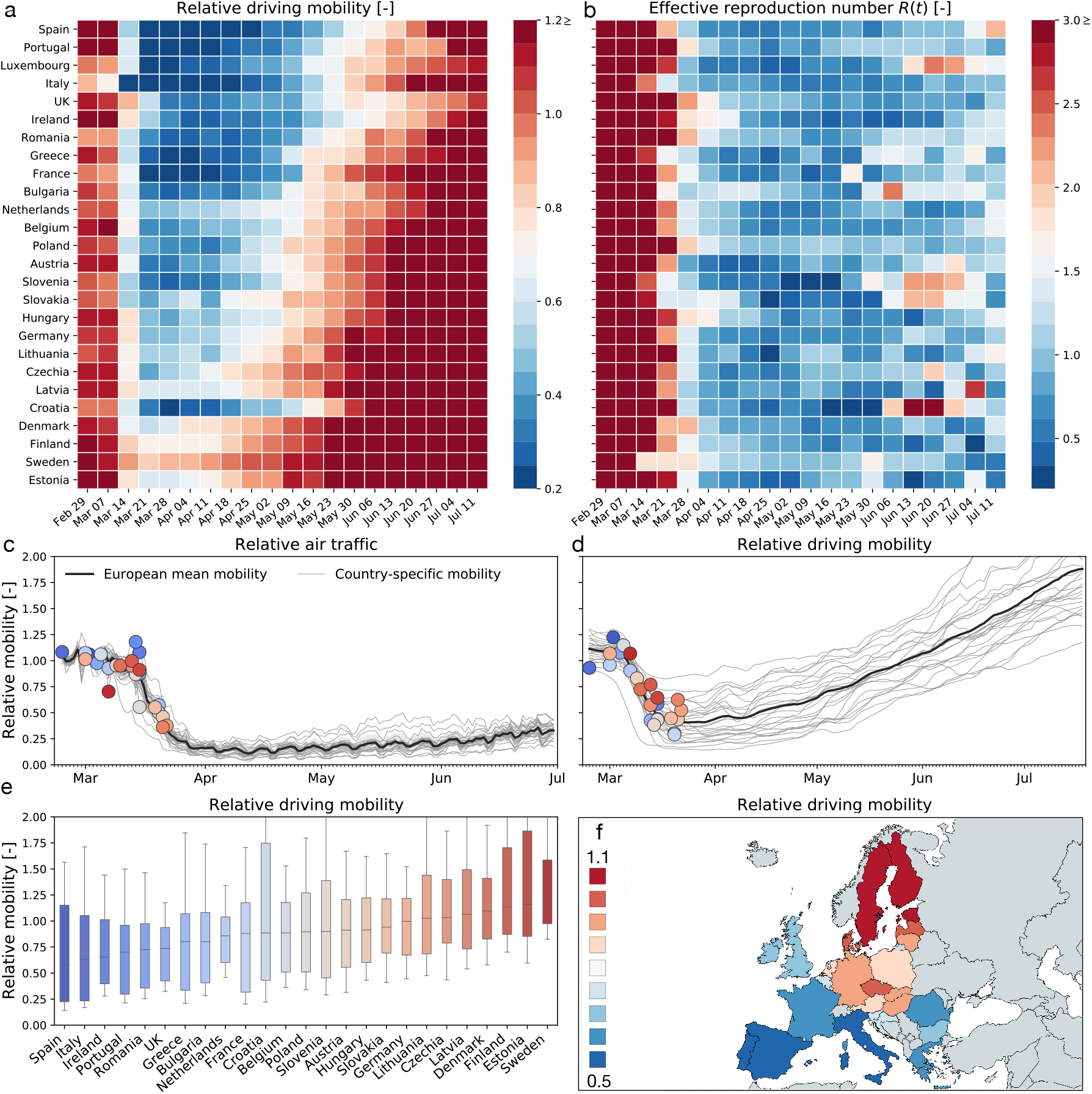
Global and local mobility across Europe. (a) weekly average relative driving mobility; (b) weekly average effective reproduction number; (c) relative air traffic; (d) relative driving mobility; (e) box plot of country-specific relative driving mobility; (f) contour plot of country-specific relative driving mobility. All mobility values are normalized with respect to baseline in early February; dots represent time point beyond which the number of cases exceeded 100, color code indicates relative mobility.

### Increased mobility only very gradually increase the effective reproduction number

Figures 2(a & b) show, side by side, the weekly average of the effective reproduction number and the relative driving mobility. At the early phase of the outbreak, this heatmap shows that a reduction of driving mobility is followed by a reduction of the effective reproduction number with a typical delay of about two weeks. At the later phases, however, an increase in driving mobility in response to a gradual exit from lockdown does not initiate an equally steep increase in the effective reproduction number. This lack of symmetry between reduction and increase in mobility suggests that some other non-pharmaceutical interventions that were adopted in that period may have taken a more prominent role in controlling the effective reproduction number.

### Local mobility is highly correlated with the reproduction number

Following the suggestion of Fig. 2(a & b) that there is an association between mobility and disease dynamics, we show in Fig. 3 the evolution of both local mobility and effective reproduction number for all countries that had at least 100 cases on March 10. To ensure similar initial conditions, we begin each simulation at the individual date the country hit 100 confirmed cases. We compute the effective reproduction number, *R*(*t*), by the semi-parametric Gaussian process model and use only the country-specific mobility data over time as input. We train the Gaussian process model in a hierarchical manner and share the priors for the model between all countries. The close agreement of the fit in Fig. 3 indicates that the model is capable of learning the individual behavior based on shared posteriors. The hierarchical posterior distribution of the adaption time *t^*^* in Fig. 3 shows a mean response time of 18.8 ± 2.2 days, with regional variations ranging from 11.2 days in Austria to 26.7 days in Sweden. We also determined the associated cross-correlation between the local mobility and the effective reproduction number. The time lag Δt varies from the shortest time interval with 14.6 ± 5.6 days to the longest interval with 30.3 ± 6.3 days. Fig 4(a) suggests a gradual loss of correlation between reproduction and mobility with increasing time interval length. The ten graphs of the ten countries compare the correlation versus time lag for six different time intervals, all starting on March 1, and 72, 87, 102, 117, 132, 147 days long, from red to blue, as highlighted in Fig 4(d). Increasing the time interval decreases the correlation and increases the time lag Δt, indicated by the peak of the individual curves. Fig. 4(b) summarizes these results across all ten countries as box plots for all six intervals. Fig. 4(c) shows a similar box plot, but now for three intervals as defined by the effective reproduction number, the first wave with *R*(*t*) ≥ 1, the trough with *R*(*t*) < 1, and the second wave with *R*(*t*) ≥ 1. Fig. 4(d) shows the dynamics of the relative infectious population for each country, and, on top, the six different interval lengths used for the analysis in Fig. 4(a & b). All ten countries display a similar strong correlation between the effective reproduction number and the local mobility, albeit with varying time lags Δ*t*. Strikingly, with increasing time, this correlation decreases, i.e., the blue curves have lower peaks than the red curves, and the time lag increases, i.e., the peaks of the blue curve are further to the right than those of the red curves.

**Figure 3:**
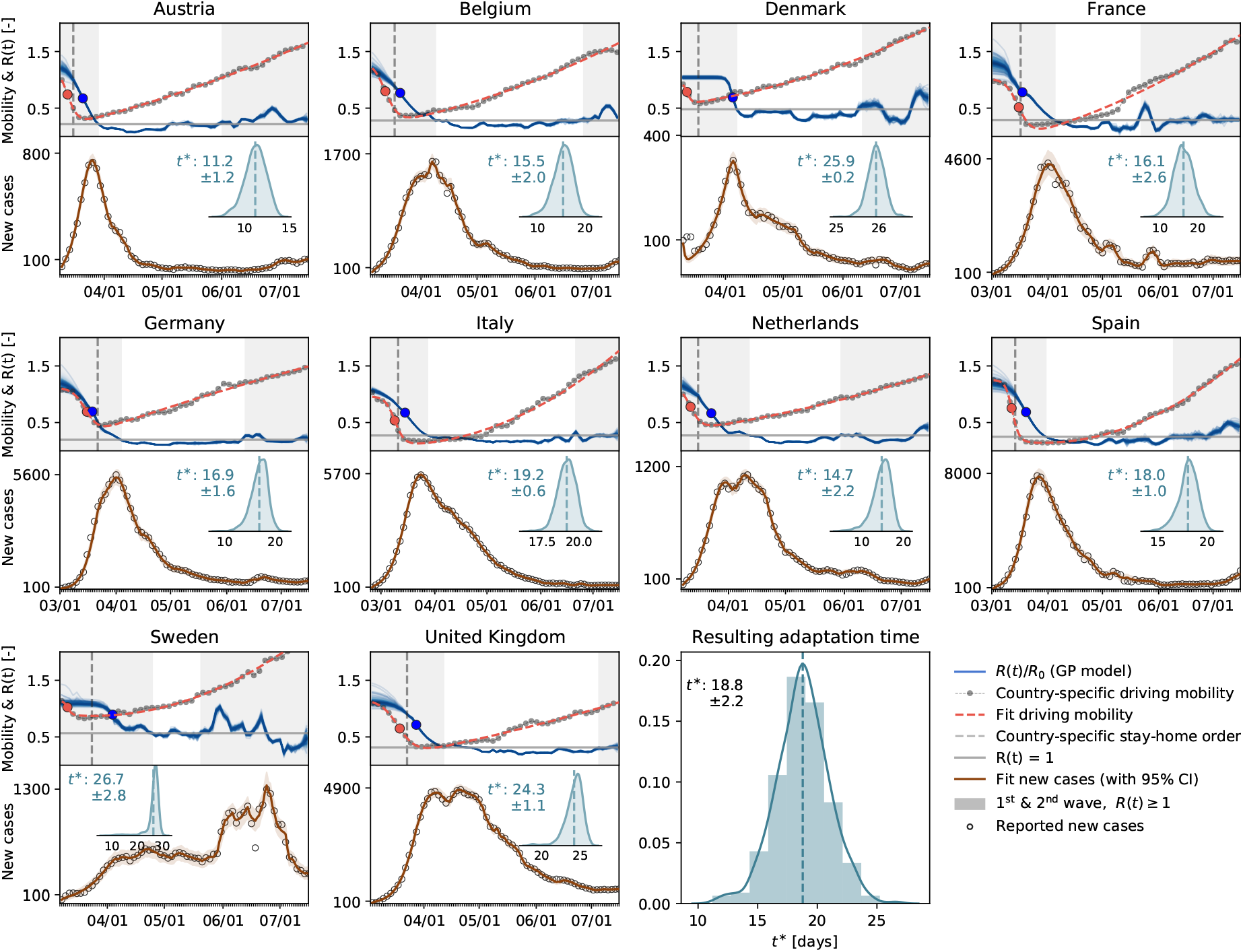
Local mobility, reproduction number, and reported cases across Europe. The hierarchical model learns the time-varying effective reproduction number *R*(*t*) (blue curve) from the reported cases (black circles) and simulated new cases (orange curve) for varying adaptation times *t^*^*. Dots in the top plots indicate the adaptations times t* of local mobility (red dot) and reproduction (blue dot)); vertical dashed lines highlight the date of stay-home order; gray regions indicate first and second waves with *R*(*t*) ≥ l, white band in between indicates the wave trough with *R*(*t*) *<* 1.

**Figure 4:**
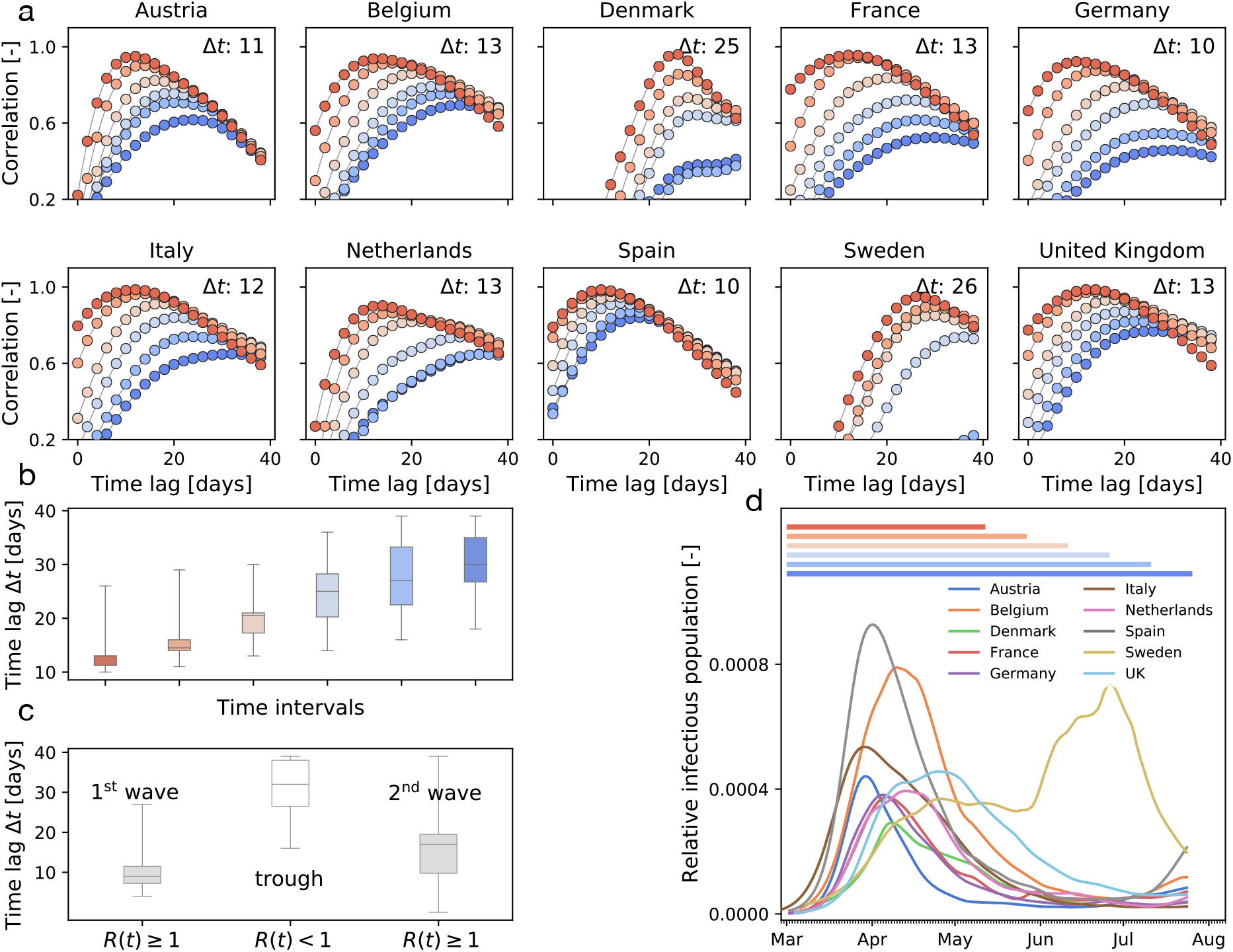
Cross-correlation between effective reproduction and local mobility. (a) correlation vs. time lag for six intervals all starting on March 1, and ranging from 72 days (red) to 147 days (blue); Δ*t* is the time lag with the highest correlation, i.e., the peak of the highest curve; (b) box plot of time lags for six intervals; (c) box plot of time lags for first and second wave with *R*(*t*) ≥ 1 and wave trough with *R*(*t*) *<* 1; (d) six time intervals for correlation analysis and relative infectious population in all ten countries.

### Mobility and reproduction number define pandemic staging

We can now combine both global and local mobility data together with the effective reproduction number to define typical staging dynamics. In Fig. 1, we use a network model for global mobility coupled with the local mobility data at each node. Following the disease outbreak in phase I, the first transition occurs following a drop in local mobility associated with changes in behavior such as working at home or limit on gatherings, and corresponds to an inflection point in the local mobility curve. In most countries, governmental outbreak control and some level of lockdown were established during the following phase II as indicated through the vertical dashed lines in Fig. 3, see also Fig. 1(c). The second transition occurs when, similarly, air traffic drops with its corresponding inflection point. During phase III, there is a slow increase in local mobility associated with a decrease in the overall reproduction number, which is still larger than one *R*(*t*) ≥ 1. The third transition between phases III and IV marks the end of the first wave and corresponds to the peak of the number of new cases. After this transition, in phase IV, the reproduction number remains consistently smaller than one, *R*(*t*) *<* 1, despite a gradual increase in mobility, resulting in a noticeable reduction of new active cases. Phase IV marks the trough, the valley between the first and second waves. As all countries gradually release their control measures, the increasing mobility will ultimately trigger an increase of new cases beyond a point, where the effective reproduction number will again become larger than one, *R*(*t*) ≥ 1. This marks the beginning of the second wave. Fig. 3 illustrates the first and second waves with *R*(*t*) ≥ 1 in grey and the wave trough with *R*(*t*) *<* 1 in white and Fig. 4 (c) illustrates the clear distinction of the time lag Δ*t* during these three intervals.

### Mobility data can be used to forecast reproduction

The observed high correlation between mobility data and the effective reproduction number, with a typical delay of around 14 days, suggests that we can use mobility data as a barometer for the reproduction number. In Fig. 5 we show two different training scenarios where we tested the prediction for the 14 days from April 19 to May 2 for Germany and the United Kingdom. In the first scenario, we used data from February 28 until April 19 for training. This period includes a rapid change in car traffic and the implementation of several different control strategies. The second period starts at the point where local mobility is close to a minimum in Europe, therefore we included case data after March 25 into the training dataset. We observe that, although a smaller training set is used, the model prediction for the effective reproduction number is better and fully captures the reproduction dynamics. Figure S3 demonstrates this predictive capabilities of the model for all ten countries.

**Figure 5:**
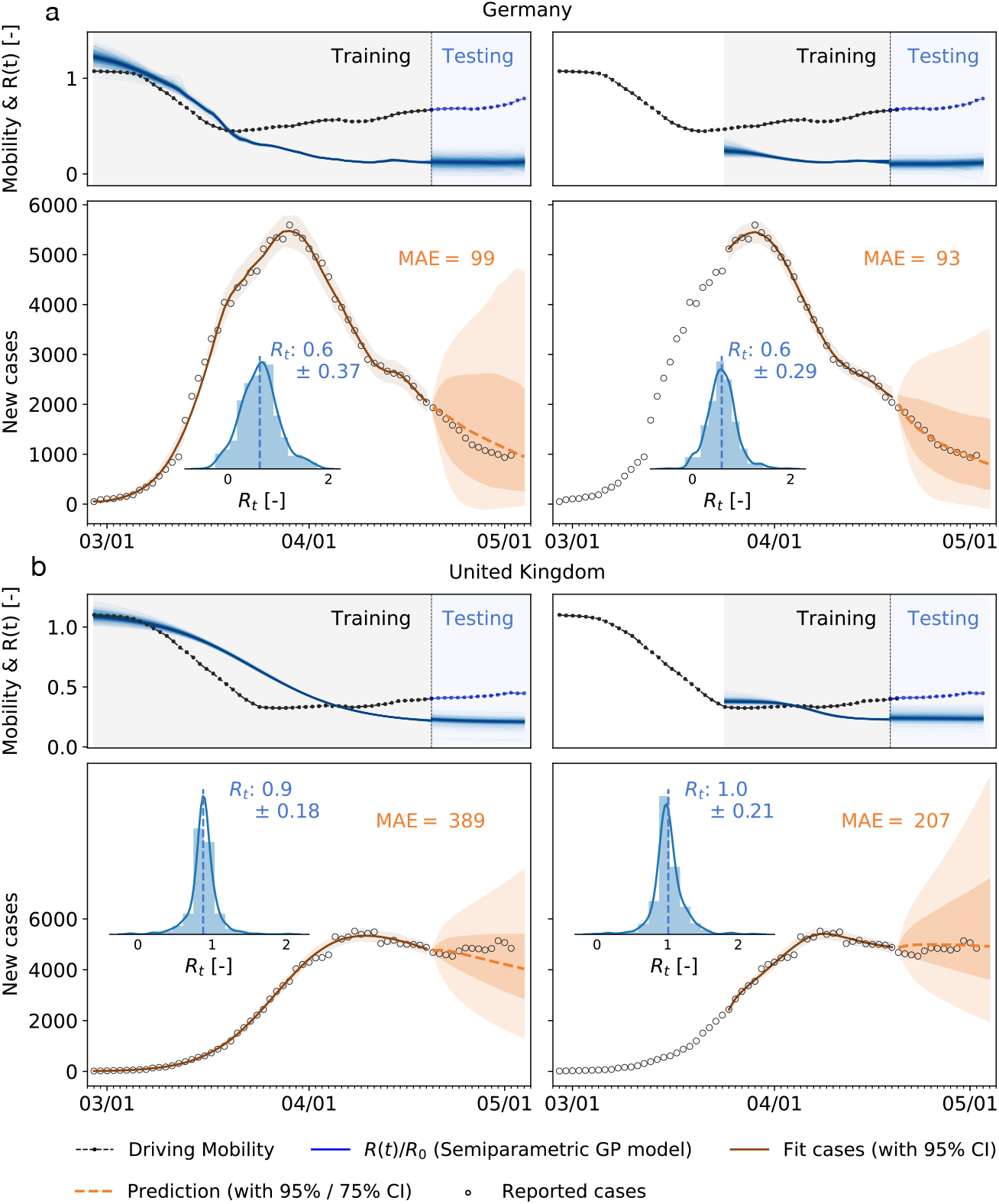
Local mobility, reproduction number, reported cases, and forecasting for Germany and the United Kingdom. Both models are trained on the full data set and on the data points after the mobility break down. Histograms show posterior distributions of last time step of predicted effective reproduction number *R*(*t*). Orange dashed curve indicates the 14-day forecast, orange zones indicate the 95% and 75% confidence intervals and associated mean absolute errors.

### Local mobility data can be used to identify possible super-spreading events

Super-spreading events are characterized by a local outbreak when a large population comes in close contact for a significant period of time. Examples includes sporting events, parades, festivals, religious ceremonies, and carnivals. Unavoidably, these events are associated with large population movement and can be captured by mobility data. For instance, the Carnival season between February 20 and 26 in the Heinsberg district drew large crowds and it is believed to be the first COVID-19 hotspot in Germany (*16*). Figure 6 shows the mobility data of the city of Cologne, approximately 70km from Heinsberg. The spike in mobility is directly followed by the epidemic outbreak and correlated with a change in the reproduction number with a adaptation time of 17 days. This highly-localized event rippled across the entire country with a adaptation time of about 40 days.

**Figure 6:**
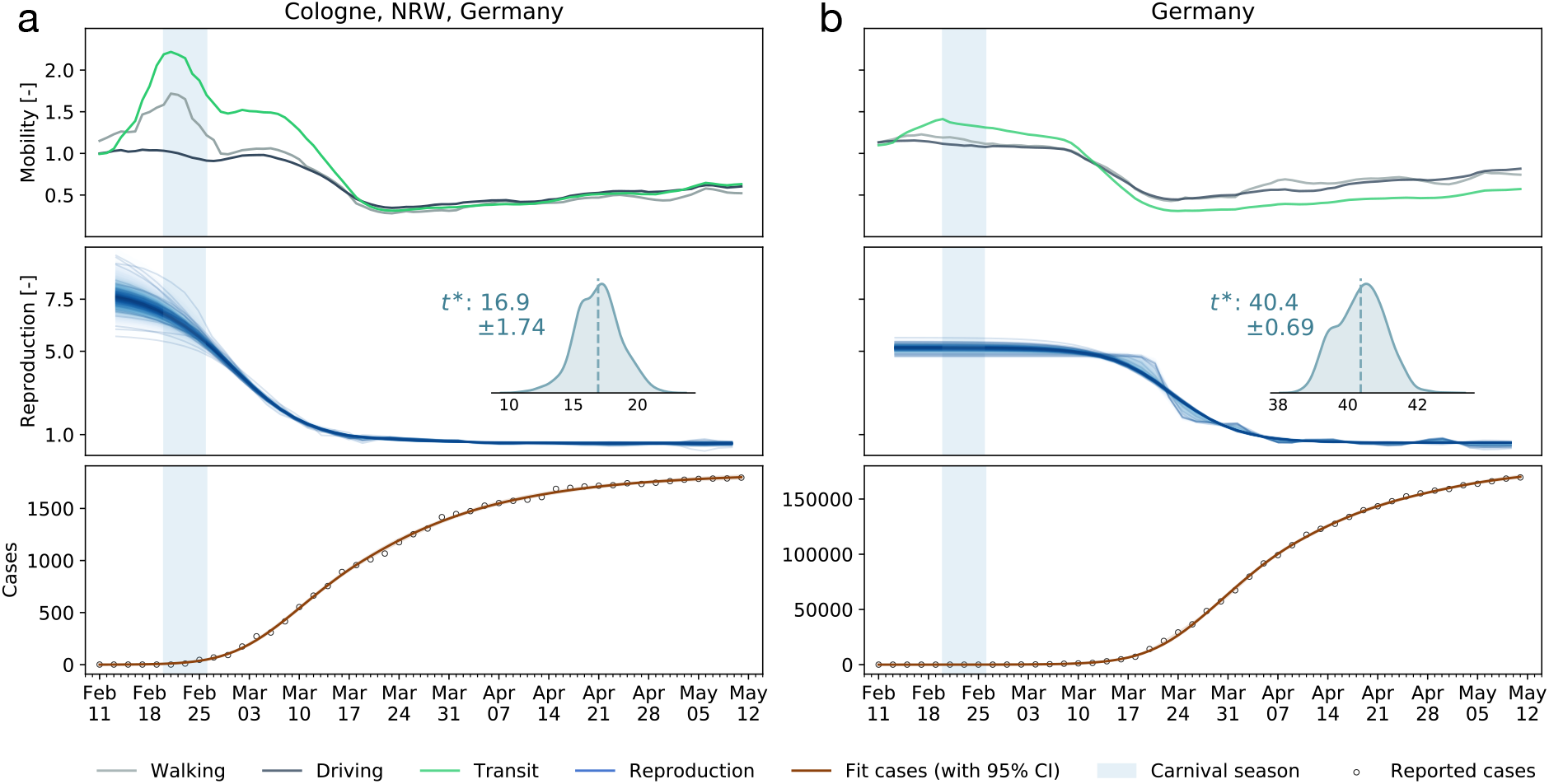
Dynamics of a super-spreading event. (a) The Heinsberg Carnival took place between February 20 and 26 (shaded region) and is associated with a large spike in mobility data in the nearby city of Cologne. (b) It is correlated with a local increase of the reproduction number in Cologne, approximately 17 days delayed, and a national increase, approximately 40 days delayed. Cases are depicted as cumulative case numbers.

## Discussion

The objective of this study was to explore to which extent global and local mobility are correlated with the effective reproduction number, and, accordingly, with the local outbreak dynamics. Using passenger air travel, cell phone mobility data, and reported COVID-19 cases across Europe, we showed that mobility and reproduction are correlated during the early stages of the outbreak, but become decorrelated during later stages. Interestingly, our study identified four distinct phases of the outbreak across all European countries that implemented political counter measures.

### Phase 0: During the early stages of the pandemic, global mobility modulates the initial outbreak pattern

Various studies have shown that, especially during the very early stages of an outbreak, there is a close correlation between mobility and spreading of an infectious disease. For instance, the early pattern of COVID-19 closely mimics passenger air travel (*17*). Global mobility is key to seed the disease in new locations before its local growth. Tight travel restrictions and border control, first implemented in the United States, and then in the entire European Union, mark the end of this phase in Mid March. In response, air travel within the European Union dropped by 95% within less than two weeks. Yet, it is becoming increasingly clear now that most travel restrictions were implemented too late to protect any country from a local outbreak of COVID-19 (*6*). While global mobility as in Fig. S4 can be an important modulator at very low or zero case numbers (*18*), our study suggests that a country-specific analysis based on local mobility alone as in Fig. 3 is probably sufficient to explain the major outbreak dynamics. In addition the current study focuses only on mobility, while other factors, for example, self-adopted human behavioral changes, could show similar correlation patterns.

### Phase I: Once a location is hit by the pandemic, exponential growth governs the local outbreak dynamics

After local seeding, the outbreak dynamics become decorrelated from global mobility. Instead, the local number of cases increases rapidly and the question of health care resources becomes the focal point in political decision making. At this point, without any additional measures, the outbreak would naturally peak and decay towards the endemic equilibrium (*26*). The timing of the peak, its magnitude, and condition of herd immunity are determined by the basic reproduction number (*20*). For COVID-19, the basic reproduction number *R*_0_ is on the order of three to six (*19*), for which models would predict a peak of active cases from 21% to 39% of the population, occurring between days 46 and 23, and herd immunity after 67% to 86% of the population have become infected.

### Phase II: Outbreak control modulates the effective reproduction number by reducing local mobility

To stop the period of exponential growth, political measures, including local lockdown and limit to gatherings, have been implemented in almost all European countries to limit contact between infectious and susceptible individuals. The rapid reduction of car traffic to 40±21% within less than two weeks in early March is an indicator for a successful contact reduction. A clear limitation of our study is the use of Apple mobility data (*23*) as a proxy for the local mobility of individuals within a country. A comparison with alternative provider-based mobility data (*25*) shows that, at least for the example of Germany, the Apple data overestimate the true reduction in mobility by up to 25%. This suggests that the Apple data are biased towards a subset of the population that can potentially respond more rapidly and more flexibly to the new stay-at-home conditions. While we clearly have to be careful to draw conclusions from mobility in absolute numbers, we believe that the general trends are indicative of a universal reduction in mobility within the general population. Strikingly, in most countries, this reduction emerged *naturally*, well ahead of political intervention, as a result of voluntary behavioral changes in the population, see Fig. 2.

### Phase III: Reduced mobility reduces the number of new cases and initiates a flattening of the curve

In the outbreak dynamics, reduced local mobility induces a reduction of the effective reproduction number *R_t_* and with it convergence to an enforced equilibrium state, a converged state under given constraints, long before herd immunity is achieved in the entire population. The speed and magnitude by which the reproduction number drops are a measure of the effectiveness of public health interventions. For example, in Austria, a country that is known for its strict response to the pandemic *R_t_* dropped from 4.0 to 1.1 in only 16 days; in Sweden, a country that implemented relatively lose public restrictions it dropped from 2.0 to 1.1 in as much as 33 days. A limitation of our study is its sensitivity with respect to the date of reporting, which can vary significantly between the different countries. We illustrate the impact of drawing the data from two different days of reporting, the onset of symptoms and the reported test results, and show the resulting time shift for the example of Germany in Fig. S2. In the hypothetical case of complete lockdown, the number of new cases, and with it the effective reproduction number, would go to zero. However, Fig. 1(c) shows that even under the strict political constraints imposed in most European countries, the average duration from the point a country hit 100 cases until the reproduction number went down to one is 38 ± 8 days. As long as *R_t_* remains lower than one, the number of new cases continues to decrease, although gradually.

### Phase IV: The gradual exit from lockdown reveals the effect of social learning

Following a period of rigorous lockdown, most European governments have begun to slowly relax the strict measures to reduce movement and contacts. This relaxation is clearly reflected in the mobility data in Fig. 2 as a slow increase of local mobility. Two important features are now becoming apparent: First, there is an asymmetry in the decrease and increase of the reproduction number before and after lockdown that can be attributed to social learning and soft political intervention including face masks, physical distancing, and no large gatherings. Second, variations in the reproduction number during the early phases are highly correlated with changes in local mobility, while later they are not. Our analysis with training and testing suggests that, as long as the disease remains endemic, *trends in local mobility allow us to forecast the outbreak dynamics for a two weeks window*. This correlation has potentially important consequences: Some proposed exit strategies suggest either periodic or on-demand lockdown once the number of new cases begins to rise again. The general objective of most outbreak control strategies is to keep the effective reproduction number either strictly below one or slightly above. The problem with the latter approach is that by the time a critical rise becomes observable, the inertia of the exponential dynamics is already difficult to control and long periods of lockdown would become necessary to manage the overall outbreak dynamics. Our study shows that mobility data can serve as a barometer to adjust particular sectors of the economy, in real time, to maintain *future* effective reproduction numbers smaller than one. Importantly, in this approach, the forecast of the reproduction number, the correlation levels, and the window of correlation can be updated daily as new data become available.

### 2nd Wave: Complete decorrelation between mobility and reproduction indicates the end of the first wave

In the case of COVID-19, during the early stages of exponential growth, almost all European countries experienced a strong reduction in local mobility. Around Mid March, numerous political measures were implemented in different stages, ranging from prohibiting gatherings to complete lockdown, with a restriction to essential business only. During this period, the virus spread non-linearly across Europe, see Fig. 3 and Fig. 4(c). Throughout this time window, individual mobility had a massive potential to accelerate the outbreak and generate large effective reproduction numbers. This agrees well with our observation of a distinct time lag and associated strong correlations between mobility and reproduction among all European countries within the full dataset, see Fig. 4(a). Due to behavioral changes after the first phase of the outbreak, our analysis suggests that the time lag increased from initially 14.6 ± 5.6 to 30.3 ± 6.3 days, before mobility and reproduction decorrelate. This is clearly visible in the cross-correlation for different time intervals of the dataset, see Fig. 4(a & b). The decorrelation begins after the infectious curve peaked, i.e., *R*(*t*) *≤* 1, and the dynamics of traffic and reproduction become increasingly independent as the effective reproduction approaches zero. In this range, the amount of infectious individuals is too low to be amplified by human mobility. For example, for the case of Austria, which was among the first countries in Europe to aggressively implement outbreak control, we observe an early peak at the end of April in Fig. 4(c). Austria was one of the first countries worldwide to report an effective reproduction number below one, which resulted in an early decorrelation and a vanishing distinct time shift between local mobility and reproduction, see Fig 4(a). The effect of decorrelation at low new case numbers is inherently built into our model: The latent variable formulation of the effective reproduction number *R*(*t*), as a function of local mobility within the SEIR model, become increasingly less sensitive to mobility changes for a low infectious population. This trend is robust and also holds for more complex compartment models including the SEIIRD model with an undetected population of asymptomatic individuals and a simultaneously fitted deceased population as we show in Fig. S1. Interestingly, the new increase of the effective reproduction number beyond one, *R*(*t*) ≥ 1, in response to the gradual exit marks a new increase in cases and the beginning of the second wave, that we can clearly see in Figs. 1 and 3.

Taken together, the strong correlation between mobility and reproduction in the endemic phase, and the loss of correlation in the later phases, suggest that mobility is a reliable predictor of to predict outbreak dynamics. Our study suggests that biweekly mobility trends can be used to stratify different phases of the pandemic, identify super-spreading events, and guide political decision making. Mobility is a suitable pandemic barometer.

## Methods

The foundation of our modeling methodology, to account for human activity at different scales, is the standard Susceptible - Exposed - Infectious - Recovered SEIR compartment model. We model the inter-country specific outbreak dynamics using a network of passenger air travel between individual countries, combined with the local driving mobility at the nodal level. To account for our prior knowledge of the model parameters and the available data for each country, we combine our network model with a hierarchical Bayesian parameter inference by applying Markov Chain Monte Carlo sampling to obtain posterior distributions for uncertainty quantification.

### COVID-19 outbreak data, global and local mobility

We draw the COVID-19 outbreak data for 26 European countries from the reported confirmed cases from February 24, 2020 (*21*). From these data, we extract the new reported cases, Δ*Î*(*t*) = I(*t*_n+1_) − *I*(*t*_n_), as the difference between today’s and yesterday’s reported cases (*18*). To estimate global mobility, we use the passenger air travel data between any two European countries (*22*), normalized by the baseline mean air travel volume from 15 January to 15 February 2020 (*24*). To approximate local mobility, we use the Apple mobility data that summarizes the relative volume of location requests per country, region, subregion, or city, normalized by the baseline volume on 13 January 2020 (*23*). We smoothen the weekday-weekend fluctuations in outbreak and mobility data by applying a moving averaging window of seven days. We analyze the data from the country-specific first day of the outbreak on which 100 infected individuals are reported in each of the 26 European countries as illustrated in Fig. S4.

### Local epidemiology modeling

We model the local epidemiology of the COVID-19 outbreak using an SEIR model with four compartments, the susceptible, exposed, infectious, and recovered populations, governed by a set of ordinary differential equations (*29*),

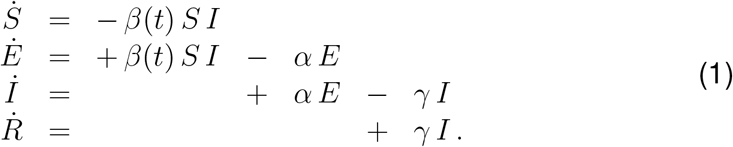

The transition rates between the four compartments, *β*(*t*), *α*, and *γ* are inverses of the contact period *β*(*t*) = 1/*β*(*t*), the latent period *A* = 1/*α*, and the infectious period *C* = 1/*γ*. We interpret the latent and infectious periods *A* and *C* as disease-specific, and the contact period *B*(*t*) as behavior specific. Using the dynamic contact period *B*(*t*), we calculate the effective reproduction number *R*(*t*) = *C/B*(*t*) as an important measure to quantify the current outbreak dynamics.

### Global network modeling

We model global mobility using European passenger air travel within a weighted undirected graph 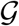 with *N* = 26 nodes and *E* = 201 edges (*17*). The nodes represent the individual countries, the edges the most traveled connections between them. We weight the edges by the estimated annual incoming and outgoing passenger air travel statistics (*24*) from which we create the adjacency matrix, *A*_ij_, that represents the travel frequency between two countries i and j, and the degree matrix,

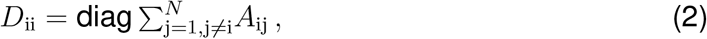

that represents the number of incoming and outgoing passengers for each country i. The difference between the degree matrix *D*_ij_ and the adjacency matrix *A*_ij_ defines the weighted graph Laplacian (*17,28*),

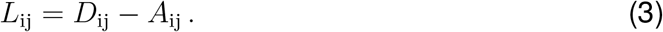

Fig. 1(b) illustrates the discrete graph 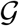 of the European Union. The size and color of the nodes represent the degree *D*_ii_, the thickness of the edges represents the adjacency *A*_ij_. For our passenger travel-weighted graph, the degree ranges from 222 million in Germany, 221 million in Spain, 162 million in France, and 153 million in Italy to just 3 million in Estonia and Slovakia, and 2 million in Slovenia, with a mean degree of 48 ± 64 million per node. To calculate the weighted Laplacian *L*_ij_, we use the air travel data across the European countries and the United Kingdom (*24*) normalize it such that its largest coefficient is equal to one, and then scale it with the air mobility coefficient *ϑ*. We discretize our SEIR model on our weighted graph 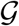 and introduce the susceptible, exposed, infectious, and recovered populations *S*_i_, *E*_i_*, I*_i_, and *R*_i_ as global unknowns at the i = 1,*…,N* nodes of the graph 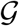. This results in the spatial discretization of the set of equations with 4 *N* unknowns,

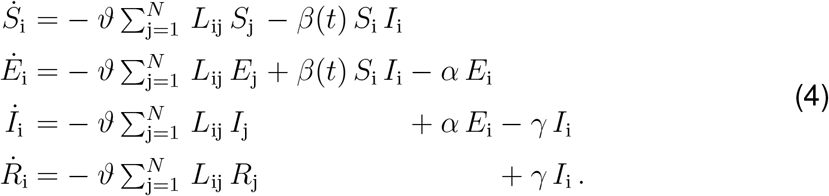

We discretize the system of ordinary differential equations (1) in time and approximate the time derivatives 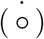 by the differences Δ(○) of the populations at two consecutive days, 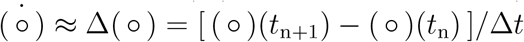, where Δ*t* =1 day.

### Human mobility modeling

It is useful to capture the general trend of mobility through simple mathematical expression so that they can be easily integrated within the epidemiological model. Here, we introduce a simple ansatz for the global and local mobility. The early phases of the outbreak are characterized by a smooth mobility transition from the initial baseline mobility to a reduced mobility induced by behavioral changes in the population. Previously, we have modeled this transition by a hyperbolic tangent-type ansatz (*29*). Here we generalize this approach by taking into account policy relaxations and social adaptations through a combination of exponentials,

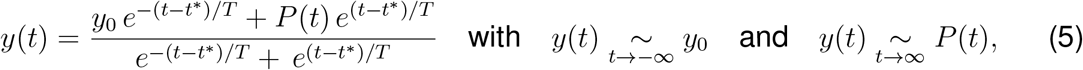

where *y*_0_ denotes the baseline value, *t^*^* is the adaptation time, *T* is the transition time, and *P*(*t*) is the adaptation after the initial drop. For the global mobility, European air traffic dropped to a constant plateau of 5% until the end of May 2020, and we select a constant *P*(*t*) to model this plateau, see Fig. 1(a). For the local mobility, driving mobility steadily increased after an initial drop at the end of March 2020, and we select a quadratic polynomial *P*(*t*) to represent this increase, see Fig. 1 (a). We assume that the the effective reproduction number follows these mobility patterns, we apply the same functional relation for the effective reproduction number,

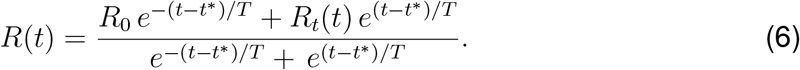

This functional form describes a smooth transition from the basic reproduction number *R*_0_ at the beginning of the outbreak to the current reproduction number *R_t_*. We model the effect of the individual mobility on the outbreak dynamics after the initial mobility drop by postulating that the current reproduction number (6) is a function of the time-varying individual mobility *R_t_* = *f* [*x*_m_(*t*)]. To keep our mobility model simple, interpretable, and capable of handling real-world data, we adopt a stochastic process approach to define *R_t_*(*x*_m_[*t*]) and construct a Gaussian process latent variable model. The Gaussian process model can be considered as a prior distribution for the mapping function (*30*),

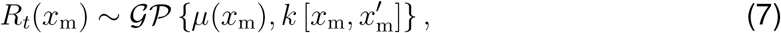

which draws function values from a multivariate normal distribution, parameterized by the mean function *μ*(*x*_m_) and the covariance function 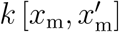, while assuming *R_t_*(*x*_m_) to be constant within a time window of two days. To account for a smooth non-linear mapping from the latent to the data space, we choose an exponentiated quadratic form covariance function with the two kernel hyperparameters *η*^2^ and *ℓ^2^* (*31,32*). A powerful way to stabilize time-series predicting models is to enable trend changes at learned time points *S*_p_ with the aid of weakly informative priors (33). These change points can be at any given time point *s_j_* with *j* = 1,2,*S*_p_. Here we apply a simple piece-wise linear trend change in the mean function (*33*),

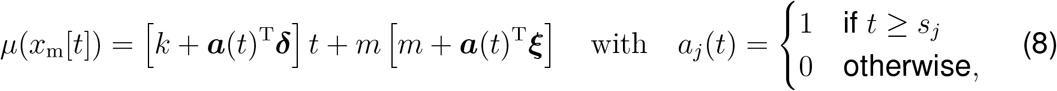

where *m* is the offset, ***δ*** is the rate adjustment, *k* is the growth rate, and *γ* enforces continuity as ξ*_j_* = *−s_j_δ_j_*.

### Bayesian parameter inference

We need to estimate a set of 12 parameters including a set of four parameters for the local SEIR model, ***θ***_SEIR_ = { *α, γ*, *E*_0_, *I*_0_}, a set of seven parameters for the semi-parameteric model, ***θ***_Rt_ = {*t^*^,T, η*^2^*, ℓ*^2^*,k,m*,***δ***}, and one parameter for the network model *ϑ*(*t*). We estimate the mobility parameter *ϑ*(*t*) by fitting Eq. (5) against the aviation data in Fig. 1(a) and multiplying it with a scaling prior *ϑ*_0_. We fix the latency period to *A* = 2.5 days and the infectious period to *C* = 6.5 days. Previous studies had reported a mean incubation period on the order of *A* = 5.0 days (*34,35,40*), with the actual infectiousness starting already *A* = 2.3 days after exposure (*27*), which is well in line with our previous findings of *A* = 2.6 days (*28*) and with the value of *A* = 2.5 days we selected here. The infectious period and varies strongly in the literature from *C* =1.5 days to *C* = 4.0 days up to distributions with a quick decline after *C* = 7.0 days (*27,34*), which motivated our current choice of *C* = 6.5 days. We estimate the remaining set of model parameters ***θ*** = ***θ***_SEIR_ ⋃ ***θ***_Rt_ ⋃ {*ϑ*_0_} using Bayesian inference with Markov Chain Monte Carlo sampling. We adopt a Student’s t-distribution for the likelihood between the new daily reported cases, Δ*Î*(*t*) = *Î*(*t*_n+1_) − *Î*(*t*_n_), and the model predictions, Δ*I*(*t*, ***θ***) = *I*(*t*_n+1_, ***θ***) − *I*(*t_n_*, ***θ***), with a new-case-number-dependent width (*36,37*),

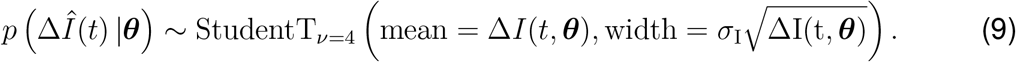

Here, *σ*_1_ represents the width of the likelihood *p*(Δ*Î*(*t*) | ***θ***) between the new daily reported cases Δ*Î*(*t*) and the modeled cases Δ*I*(*t*,***θ***). We apply Bayes’ rule to obtain the posterior distribution of the parameters on the basis of the prior distributions specified in Table S2, and the reported cases themselves,

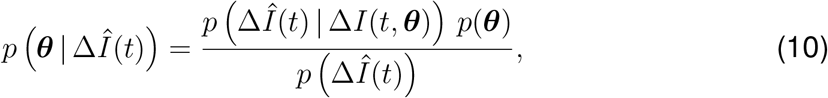

which we infer approximately by employing the NO-U-Turn sampler (NUTS) (*38*) implementation of the Python package PyMC3 (*39*). To account for variability between the individual countries, while simultaneously taking advantage of the entire dataset, we adopt a hierarchical model to learn the effective reproduction number on the basis of the case and mobility data. We postulate that during the initial phase, the effective reproduction number is primarily governed by the local political action in each country, while during the later phases, it becomes strongly correlated with local mobility that mimics the new levels of social awareness. We apply hierarchical priors on the parameters of the initial phase, i.e., the basic reproduction number *R*_0_, and the adaptation and transition times *t*^*^ and *T*, and define shared priors on the semi-parametric Gaussian process latent model of the later phases. For each county *i*, we draw *R*_0_*_i_* and 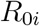 from normal distributions, 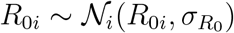 and 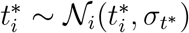, and *T_i_* from a log-normal distribution, 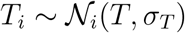, and model the hyper standard deviation priors by half-normal distributions. We design the Gaussian process latent variable model with shared priors for all countries and inform it by the time-varying local mobility data. Here we make use of two hyperparameters for the kernel functions *η*^2^ and *ℓ*^2^. For the forecasting models, we construct a linear-piece-wise means functions defined by three parameters drawn from normal distributions to stabilize the predictive capabilities. We select three equidistant change points, *S*_p_ = 3, for the piece-wise linear function. Rather than using a change point detection to identify the number and location of change points, we select three equidistant points to prevent overfitting and keep the prior distributions at a moderate level and to maintain flexibility in adjusting the change rate by using a sparse prior on the rate adjustment ***δ***. For the network model, we include one additional weakly informative prior for the scaling of the travel coefficient as a normal distribution 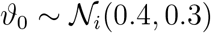. Table S2 summarizes all priors on the model parameters.

## Data Availability

n/a

## Acknowledgements

This work was supported by a DAAD Fellowship to Kevin Linka, by the Engineering and Physical Sciences Research Council grant EP/R020205/1 to Alain Goriely, and by a Stanford Bio-X IIP seed grant and the National Institutes of Health Grant U01 HL119578 to Ellen Kuhl. This work was also undertaken, in part, as a contribution to the Rapid Assistance in Modelling the Pandemic RAMP initiative, coordinated by the Royal Society for which Alain Goriely is a task leader.

## Supplementary Material

### Local epidemiology modeling - SEIIRD model

To illustrate that our method is robust to disease parameters beyond the classical SEIR model, we expand the SEIR model for the local epidemiology of COVID-19 from Eq. (1) to an SEIIRD model with six compartments, the susceptible, exposed, detected infectious, hidden infectious, recovered, and deceased populations. These six compartments are governed by the following set of ordinary differential equations,

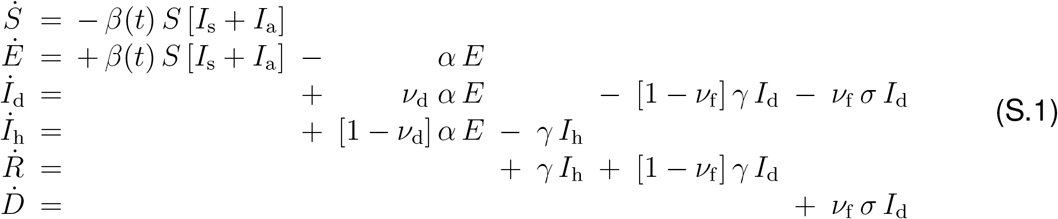

The transition rates between the compartments, *β*(*t*), *α*, γ, and *σ* are inverses of the contact period *B*(*t*) = 1/*β*(*t*), the latent period *A* = 1/*α*, the infectious period *C* = 1/*γ*, and the survival period *S*_D_ = 1/*σ*. The SEIIRD model splits the initial infectious group of the SEIR model in Eq. (1) into a detected group, *I*_d_ = *ν*_d_ *I*, and a hidden undetected group, *I*_h_ = *ν*_h_ *I*, where *ν*_d_ is the detection fraction and *ν*_h_ = 1 − *ν*_d_. From the detected group *I*_d_, individuals can transition into the recovered group *R* at a fraction [1 − *ν*_f_] or into the deceased group *D* at a fraction *ν*_f_, where *ν*_f_ is the case fatality rate. We interpret the latent, infectious and survival periods *A*, *C*, and *S*_D_ as disease-specific and static, and the contact period *B*(*t*) as behavior specific and dynamic. Using the dynamic contact period *B*(*t*), we calculate the effective reproduction number *R*(*t*) = *C/B*(*t*) to quantify the outbreak dynamics.

We need to estimate a set of 15 parameters including a set of eight parameters for the SEIIRD model, ***θ***_SEIIRD_ = { *α, γ*, *σ, ν*_d_*, ν*_f_*, E*_0_, *I*_d_,_0_, *I*_h_,_0_}, and a set of seven parameters for the semi-parametric model, ***θ***_Rt_ = {*t^*^,T, η*^2^*, ℓ*^2^*,k,m*, ***δ***}. As before, we estimate the model parameter set ***θ***^*^ = ***θ***_SEIIRD_ ∪ ***θ***_Rt_ using Bayesian inference with Markov Chain Monte Carlo sampling. We fix the latency and infectious periods to *A* = 2.5 days and *C* = 6.5 days. In contrast to the SEIR model, we now fit two data sets simultaneously, the new daily detected cases, Δ*Î*_d_(*t*) = *Î*_d_(*t*_n+1_) − *Î*_d_(*t*_n_), and the daily new deaths, 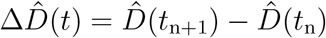, which we extract as the differences between the today’s and yesterday’s confirmed cases and deaths (*2*) and smoothen using a seven-day moving average on the data. For both fits, we adopted a Student’s t-distribution for the likelihood between the new daily reported data, Δ*Î*_d_(*t*) and 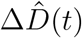, and the model predictions, Δ*I*_d_(*t*, ***θ***^*^) and Δ*D*(*t*, ***θ***^*^), with new-case- and new-deaths-number-dependent widths (*1*),

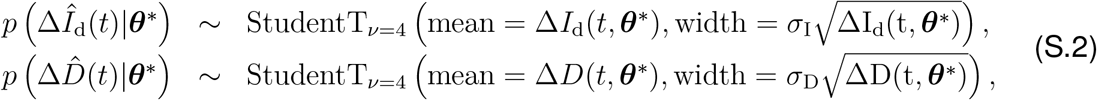

where *σ_I_* and *σ_D_* represent the widths of the likelihoods *p*(Δ*Î_d_*(*t*) |***θ***^*^) and 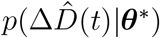 between the daily new reported cases and deaths Δ*Î*_d_ and 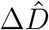 and the associated modeled new cases and deaths Δ*I*_d_ and Δ*D*. Again, we apply Bayes’ rule to obtain the posterior distributions of the parameters on the basis of the prior distributions specified in Table S3, and the reported cases and deaths themselves, which we infer approximately by employing the NO-U-Turn sampler (NUTS) (*4*) implementation of the Python package PyMC3 (*6*).

Fig. S1 shows the reported and simulated cases and deaths across Europe. The hierarchical SEIIRD model learns the time-varying effective reproduction number *R*(*t*) from both the reported cases and deaths in Fig. S1 (a) for varying adaptation times *t*^*^. The learnt survival period is *S*_D_ = 6.62 ± 0.14 days. The adaptation time distribution in Fig. S1 (b) indicates an adaptation time of *t*^*^ = 16.6 ± 2.8 days. The box plots in Figs. S1 (c & d) show the country-specific case fatality rates *ν*_f_ and detection fractions *ν*_d_. These results suggest that our method is not only applicable to the classical SEIR model but extends equally to more sophisticated models like the SEIIRD model with additional hidden and deceased compartments, and can not only fit the reported case data, but also simultaneously reported cases and deaths.

### Sensitivity with respect to date of reporting

In our main study, we interpret the daily reported case number Δ*Î*(*t*) from a central European data base (*2*) as the infectious population and fit this case number against the daily change of the infectious population Δ*I*(*t*, ***θ***) of our SEIR model. Arguably, the “date of reporting” can mean different things for different countries, and it can vary hugely between infection, symptom onset, testing, positive confirmed, and reporting. This implies that our time delay between mobility and reproduction is highly sensitive to the local testing logistics.

Table S1 illustrates difference between symptom onset and reporting for four representative countries (3). For example, in Germany, the “date of reporting” represents the date at which a sample swab is sent to laboratory testing, although the actual date at which authorities are notified will be several days later. To illustrate the difference between symptom onset and reporting, we perform the same SEIR model analysis based on two different reported data sets *Î*(*t*), symptom onset data from the Robert Koch Institute (*5*) and reporting data from European Centre for Disease Prevention and Control (*2*), which includes the data that we used throughout this study.

**Figure S1:**
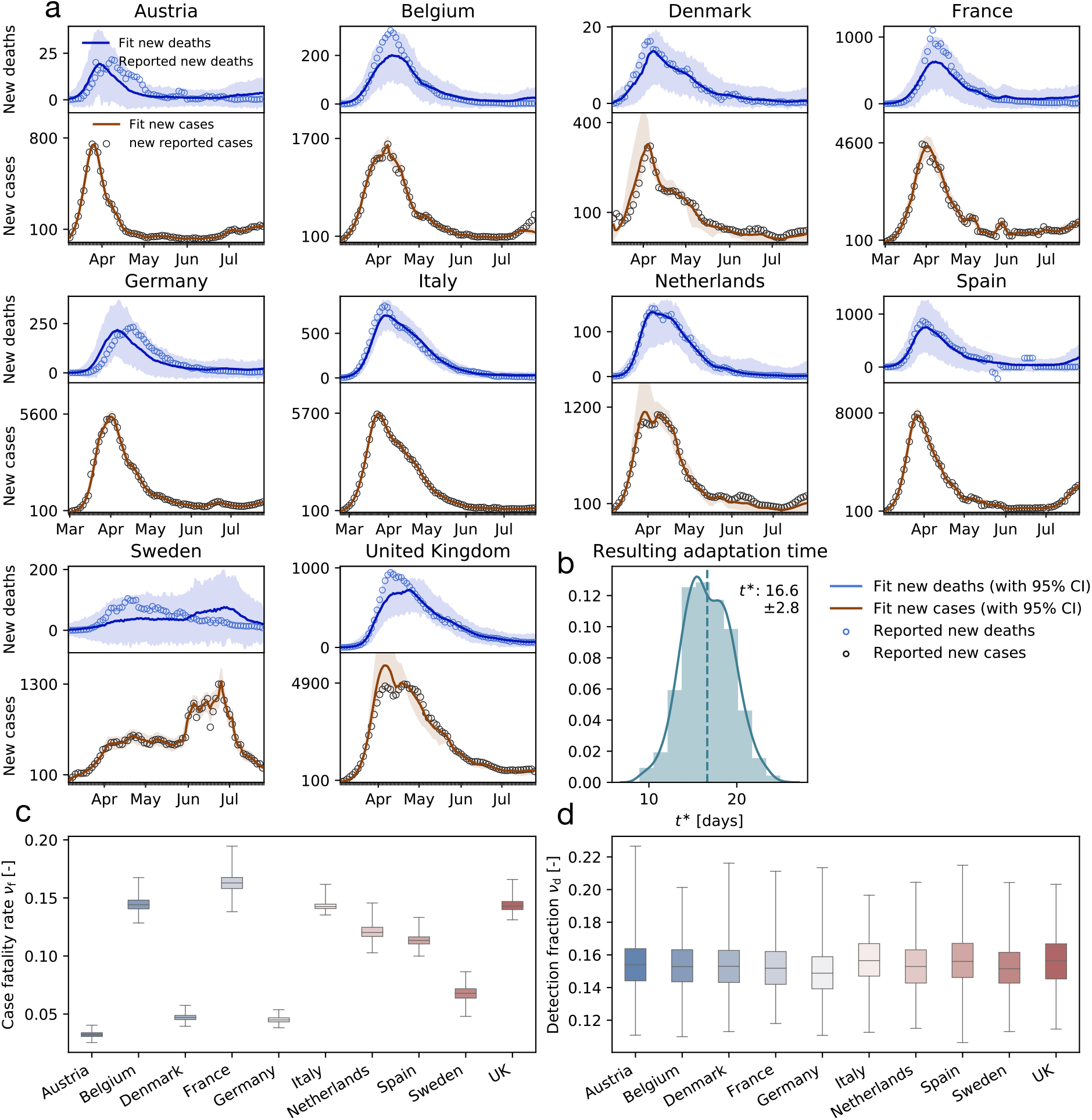
Reported and simulated cases and deaths across Europe. (a) The hierarchical SEIIRD model learns the time-varying effective reproduction number *R*(*t*) from the reported cases (black circles) and deaths (blue circles) for varying adaptation times *t*^*^; (b) adaptation time distribution indicates an adaptation time of *t*^*^ = 16.6 ± 2.8 days; (c) box plots of country-specific case fatality rates *ν*_f_; (d) box plots of country-specific detection fractions *ν*_d_.

Figure S2 illustrates the learned time-varying effective reproduction numbers for both symptom onset and reporting data, and the model fit of the simulated cases to the two data sets. The time shift in the two data sets results in a time shift of the model fit, and with it, a time shift in the effective reproduction number curves. The resulting adaptation time distributions vary between *t*^*^ = 8.10 ± 0.52 days for the system onset data and *t*^*^ = 16.80 ± 1.49 days for the reported data with a mean difference of 8.7 days. This study highlights the sensitivity of the data to the reporting logistics. Unfortunately, system onset data were not available for all countries in the European Union, and we can only show the sensitivity here, rather than using system onset data throughout our entire study, which would have been more unified and accurate.

**Table S1:**
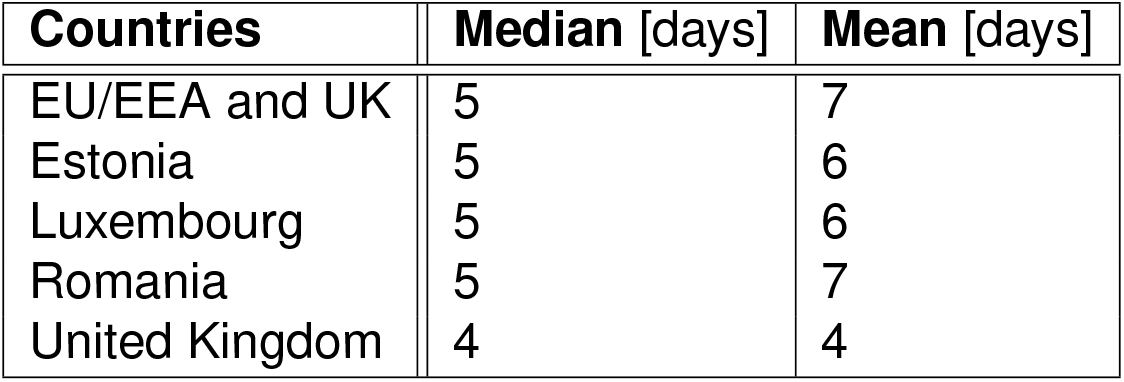
Time delay between symptoms onset and reporting (*3*)

| Countries | Median [days] | Mean [days] |
| --- | --- | --- |
| EU/EEA and UK | 5 | 7 |
| Estonia | 5 | 6 |
| Luxembourg | 5 | 6 |
| Romania | 5 | 7 |
| United Kingdom | 4 | 4 |

**Figure S2:**
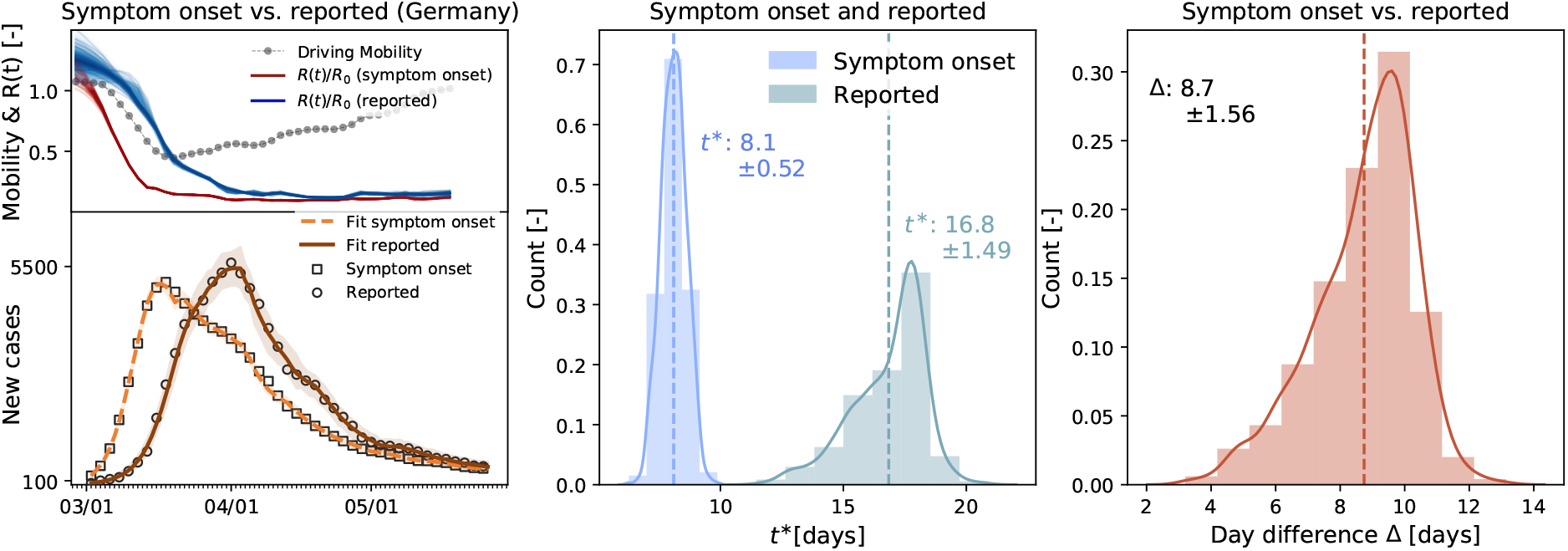
Local mobility, reproduction number, and reported cases in Germany. The model learns the time-varying effective reproduction number for symptom onset (red curve) from the symptom onset cases (black boxes) and simulated cases (orange curve) and for reporting (blue curve) from the reported cases (black circles) and simulated cases (brown curve). Distributions of adaptation time *t^*^* for symptom onset (blue) and reported data (green), and of difference between symptom onset and reported data (red).

### Forecasting

Fig. S3 compares the results of a two-week forecasting with different training sets for all ten European countries. Fig. S3 (a) uses the full available data set, whereas Fig. S3 (b) uses only a reduced data set beginning on April 1, 2020 to train the model.

**Figure S3:**
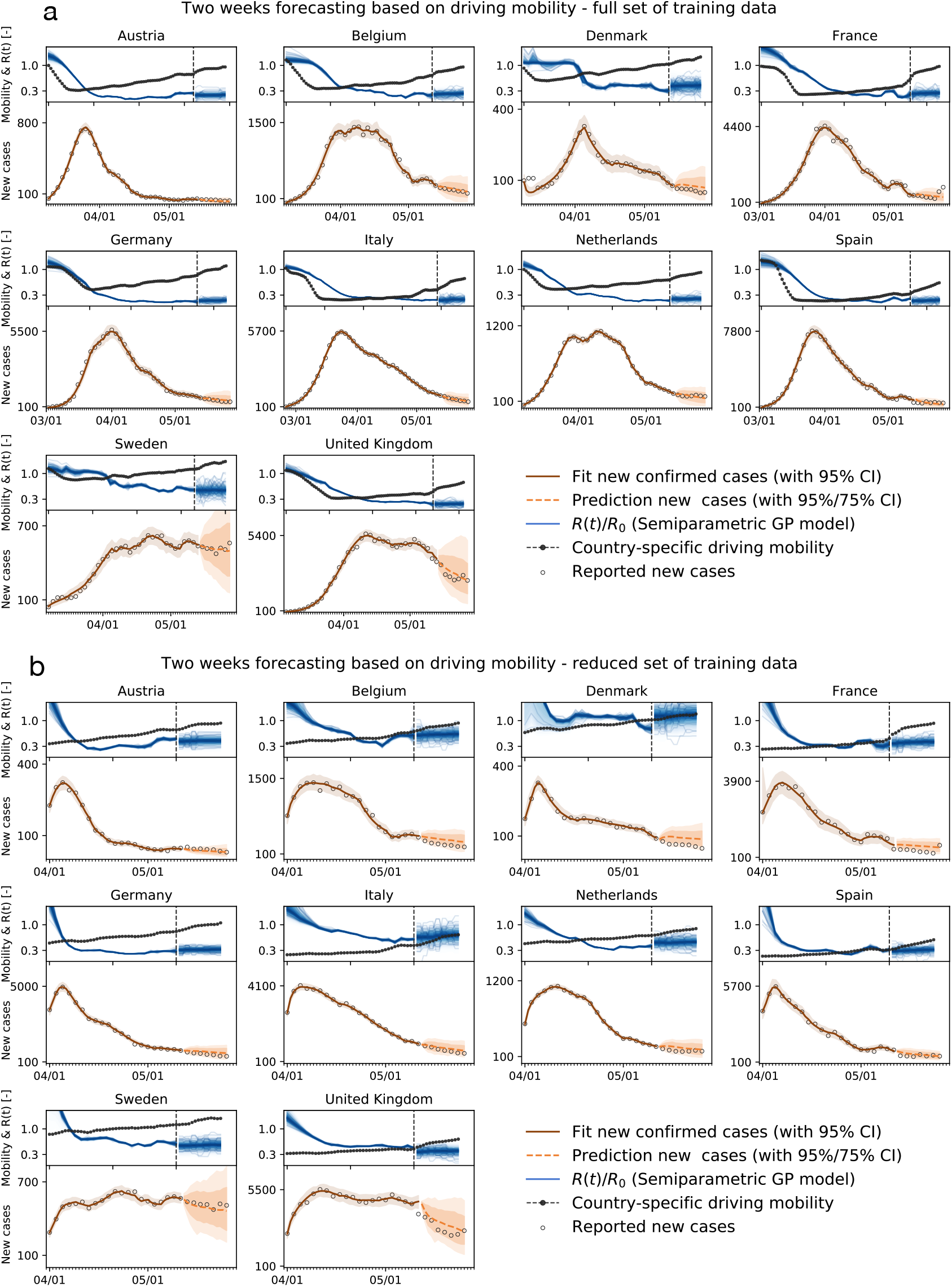
Local mobility, reproduction number, reported cases, and forecasting with different training sets. (a) Two-week forecast with training on the full available data set; and (b) Two-week forecast with training on reduced data set beginning on April 1, 2020.

### Network modeling

Fig. S4 shows results of the network model including both global and local mobility. The global mobility represents the air traffic data in each country, the local mobility represents the driving mobility.

**Figure S4:**
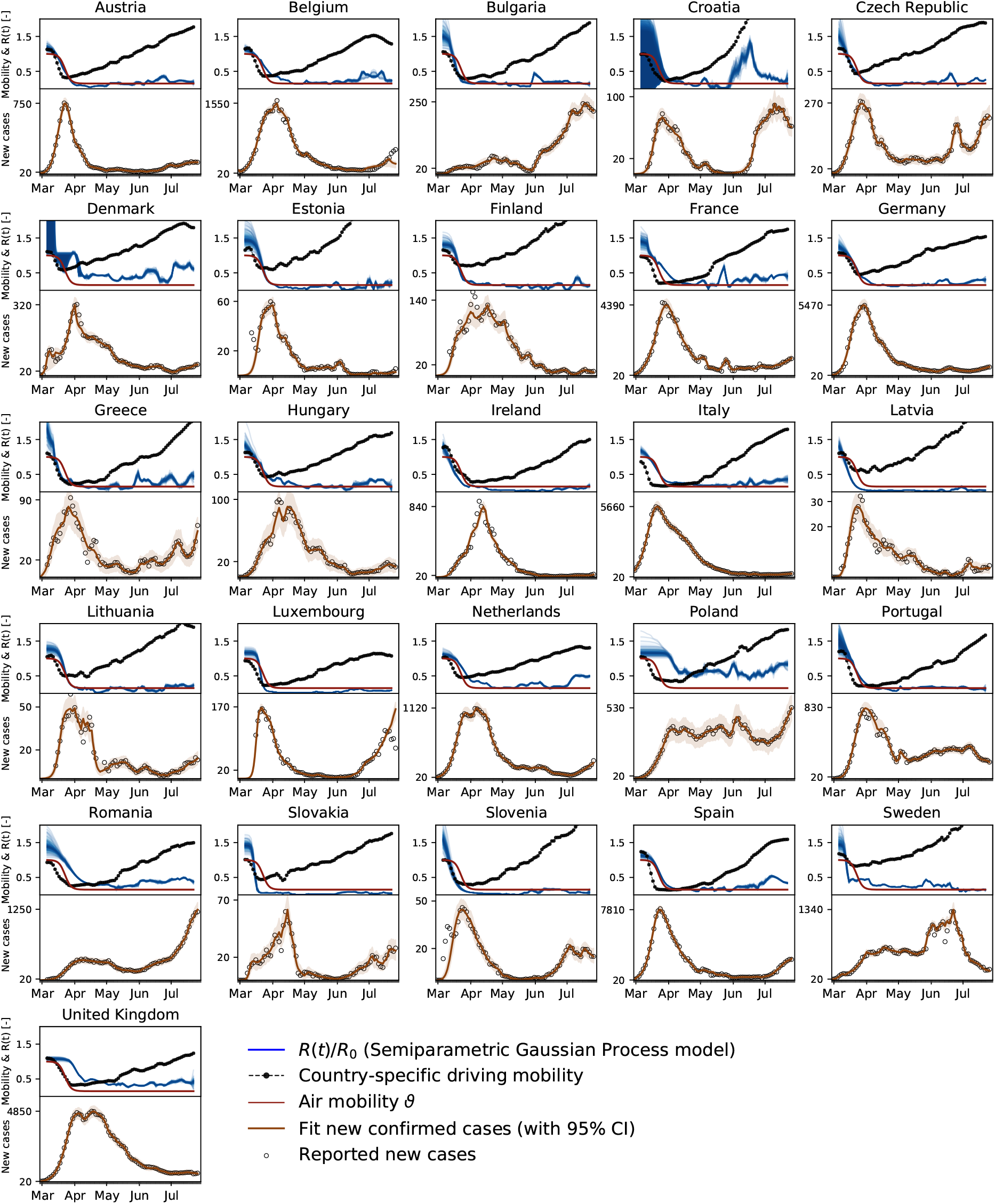
Global and local mobility, reproduction number, and reported cases across Europe using the network model. The model learns the time-varying effective reproduction number *R*(*t*) (blue curves) from the reported cases (black circles) and simulated new cases (orange curves). Global mobility (red curves) and local mobility (black curves) highlight the reduction in air traffic and driving mobility.

### Priors on the model parameters

Tables S2 and S3 summarize the priors on our the semi-parametric network model parameters and on our SEIIRD model parameters.

**Table S2:** Priors on the semi-parametric (network) model parameters.

| Parameter | Variables | Prior distributions |
| --- | --- | --- |
| $R_0$ | basic reproduction | Normal(2, 1.5) |
| $\sigma_{R_0}$ | std basic reproduction | HalfNormal(1.5) |
| $t^*$ | adaptation time | Normal(14, 14) |
| $\sigma_{t^*}$ | std adaptation time | HalfNormal(15) |
| $T$ | transition time | LogNormal($\log(3)$, 0.8) |
| $\sigma_T$ | std transition time | HalfNormal(0.8) |
| $\ell^2$ | hyperparameter | Gamma(2, 0.1) |
| $\eta$ | hyperparameter | HalfCauchy(0.5) |
| $k$ | growth rate | Normal(1, 1) |
| $m$ | offset | Normal(1, 1) |
| $\delta$ | rate adjustment | Normal(0, 2, shape=$S_p$) |
| $I_0$ | initial infected | LogNormal($\log[\Delta \hat{I}(t=0)]$, 1.0) |
| $E_0$ | initial exposed | LogNormal($\log[\Delta \hat{I}(t=3)]$, 1.0) |
| $\vartheta_0$ | travel coefficient | Normal(0.4, 0.3) |
| $\sigma_I$ | likelihood width | HalfCauchy($\beta = 1$) |

**Table S3:** Priors on the SEIIRD model parameters.

| Parameter | Variables | Prior distributions |
| --- | --- | --- |
| $R_0$ | basic reproduction | Normal(2, 1.5) |
| $\sigma_{R_0}$ | std basic reproduction | HalfNormal(1.5) |
| $t^*$ | adaptation time | Normal(14, 14) |
| $\sigma_{t^*}$ | std adaptation time | HalfNormal(15) |
| $T$ | transition time | LogNormal(log(3), 0.8) |
| $\sigma_T$ | std transition time | HalfNormal(0.8) |
| $\ell^2$ | hyperparameter | Gamma(2, 0.1) |
| $\eta$ | hyperparameter | HalfCauchy(0.5) |
| $k$ | growth rate | Normal(1, 1) |
| $m$ | offset | Normal(1, 1) |
| $\delta$ | rate adjustment | Normal(0, 2, shape=$S_p$) |
| $S_D$ | survival period | Normal(6.5, 1) |
| $\nu_d$ | detection fraction | LogNormal(log[0.15], 0.1) |
| $\nu_f$ | case fatality rate | LogNormal(log[0.01], 1.0) |
| $I_{d,0}$ | initial detected infected | LogNormal(log[$\Delta \hat{I}(t=0)$], 1.0) |
| $I_{h,0}$ | initial hidden infected | Deterministic([1 - 0.15]/0.15 $I_{d,0}$) |
| $E_0$ | initial exposed | LogNormal(log[$\Delta \hat{I}(t=3)$], 1.0) |
| $\sigma_I$ | likelihood width | HalfCauchy($\beta = 1$) |
| $\sigma_D$ | likelihood width | HalfCauchy($\beta = 1$) |

